# Mapping the flow of digital data in clinical trials and assessing its carbon footprint

**DOI:** 10.64898/2026.07.31.26358778

**Authors:** Herman Sandeep Prakasam, Neil Mackillop

## Abstract

**Background:** Healthcare contributes 5% of global carbon emissions, with clinical trials forming a meaningful share. Existing trial emission frameworks inadequately capture and account for data storage and analysis emissions.

**Objective:** (i) Map the end-to-end flow of AstraZeneca’s (AZ) trial data and identify emissions hotspots; (ii) estimate emissions from the hotspots and assess their materiality to overall clinical trial emissions.

**Design:** A top-down assessment of enterprise-level trial data volumes and associated emissions, and analysis of trial-level data-related emissions of two representative Phase III trials.

**Results:** Data flow mapping highlighted three hotspots: (a) data analysis in a statistical environment (entimICE) (b) Trial Master File (TMF) storage, and (c) short-term and long-term data storage by clinical research organisations (CROs). Within entimICE, the total volume of trial data stored for analysis across all active and recently completed trials at AZ was 100-125 TB, stored across four servers in Sweden, generating 80–100 tonnes CO_2_eq annually. TMF storage emissions fell below measurable thresholds. CROs stored substantial data volumes, but per-trial emissions were likely insignificant due to economies of scale afforded by large data centres.

**Conclusion:** To our knowledge, this is the first study to assess the GHG emissions of industry-sponsored trial data storage and analysis. It highlighted the complex network of nodes and junctions that underpin modern clinical trials. The results suggest that carbon emissions from trial data management form a small proportion and are unlikely to materially impact trial-related emissions. Future research should confirm these results in trials that employ energy-intensive operations, such as artificial intelligence (AI).

**STRENGTHS AND LIMITATIONS OF THIS STUDY:**

- This is the first comprehensive study to assess the carbon emissions from clinical trial data storage and analysis. The data flow hotspots identified in this study include (i) the statistical analysis environment, (ii) trial master file storage, and (iii) data stored by third-party clinical research organisations
- The emissions from the first two hotspots did not significantly impact overall trial emissions. CROs store large volumes of trial data, but the per-trial emissions are likely to be insignificant due to the economies of scale offered by hyperscale data centres. Overall, emissions from clinical trial data storage and analysis form a small portion of overall trial emissions.
- Energy-consuming components beyond data storage that make up a typical data server, such as backup servers, networking, and cooling systems, were not considered because isolating their functions for clinical trial data analyses was infeasible.
- The complexity of cloud data server infrastructure, confidentiality concerns regarding server infrastructure, and gaps in publicly available data made it infeasible to access numerous data points that would have improved the accuracy of the calculations. Further research must account for the emissions associated with AI.

## Introduction

Globally, 4%–5% of greenhouse gas (GHG) emissions come from the healthcare sector (<u>WHO</u>). Clinical trials are an integral component of drug development. Hence, healthcare system net-zero roadmaps are incomplete without including emissions estimates from clinical trial operations. To account for this, carbon emissions from clinical trials and the methods used to calculate them have been reported by both industry and academic groups^1,2^. The emission hotspots consistently reported in these studies include trial-related travel, blood and tissue sample management, and emissions from the trial coordination centres. These activities are therefore key focus areas to cut trial emissions^2–4^.

The Sustainable Healthcare Coalition (SHC) has developed guidance and methodology to support the estimation of carbon emissions from clinical trials^2^. Other groups have developed similar methodologies and open-source calculators^5^. However, no published framework currently exists for assessing emissions associated with the storage and analysis of clinical trial data. Given the central importance of data collection, storage, and analysis to overall clinical trial operations, the absence of such a framework is a gap that needs to be addressed to improve coverage of activity-based CO_2_ emissions in clinical trials.

The work done by Mackillop, et al. ^1^formed the foundation for this study. Similar carbon emissions from clinical trial operations and the patient pathway have been reported by others, who largely concur on the emission hotspots described above^2,3,6^. A report by Cranley et al.^7^ included a data collection and exchange module that assessed a combination of paper-based and electronic processes. The median footprint of the module across the six trials assessed in the study was 845 kgCO_2_eq (IQR 317 – 2065). The publication did not assess emissions from storing digital data on central data servers or from data analysis.

Understanding the emissions associated with clinical trial data storage and analysis is particularly relevant given the increasing integration of digital technologies in clinical trials and the growing adoption of decentralised trial designs. The digital transformation of clinical trials provides significant benefits to overall clinical trial operations, including improved efficiencies in trial execution and a reduction in trial-related travel^8^. However, increasing digital integration is linked to a larger volume of data being collected, which requires extensive data server infrastructure to store, process, and analyse it, and additional administrative steps for operational data management. Thus, the carbon impact of digitalising clinical trials must be carefully weighed.

The concerns regarding emissions from trial data storage and management are particularly significant in multicentre, global trials, where, in addition to centralised data storage, there are also localised duplicate data stores. A true estimate of the total volume of clinical trial data stored in data servers globally has not been studied and reported to date.

The importance of understanding data-related emissions in clinical trials can be appreciated by reflecting on the scale of clinical operations. Firstly, as of 2025, there were approximately 550,000 trials registered on clinicaltrials.gov^9^. Secondly, a 2022 report estimated that, on average, a Phase III study collected nearly 1 million data points. With the emergence of wearable and remote monitoring data in clinical studies, that estimate is expected to exceed 1 billion data points per Phase 3 trial today^10^. Finally, it is estimated that 55%-60% of data managed by established clinical data management CROs is stored on some form of hyperscale data server^11^.

Therefore, understanding the emissions from clinical trial data storage and analysis would: (i) help refine the activity-based carbon emissions estimation methodology for clinical trials, and (ii) inform clinical development teams on whether estimating emissions from trial data management needs attention to achieve net-zero clinical trials.

In this report, the flow of clinical trial data from the investigator site to the final TMF storage for the assessment window was mapped. Following this, the emissions associated with data storage on AstraZeneca (AZ) premises and by third-party clinical research organisations (CROs) were determined.

An overview of the flow of trial data from the investigator site to the final data repository is presented in Figure 1. For large AZ trials, numerous clinical research organisations are involved in the collection, storage, clean-up, and transmission of raw data from the investigator site to the EDC system. These vendors, shown in green boxes, collect a plethora of trial-related data and store it in their respective data server infrastructure. The trial-critical data is transmitted to the EDC system, while the vendor retains copies of the raw and cleaned data. The analysis and reporting system, i.e., the statistical analysis environment (purple box), draws data from the EDC for analysis, and the processed data is finally stored in the eTMF (yellow box).

**Figure 1.**
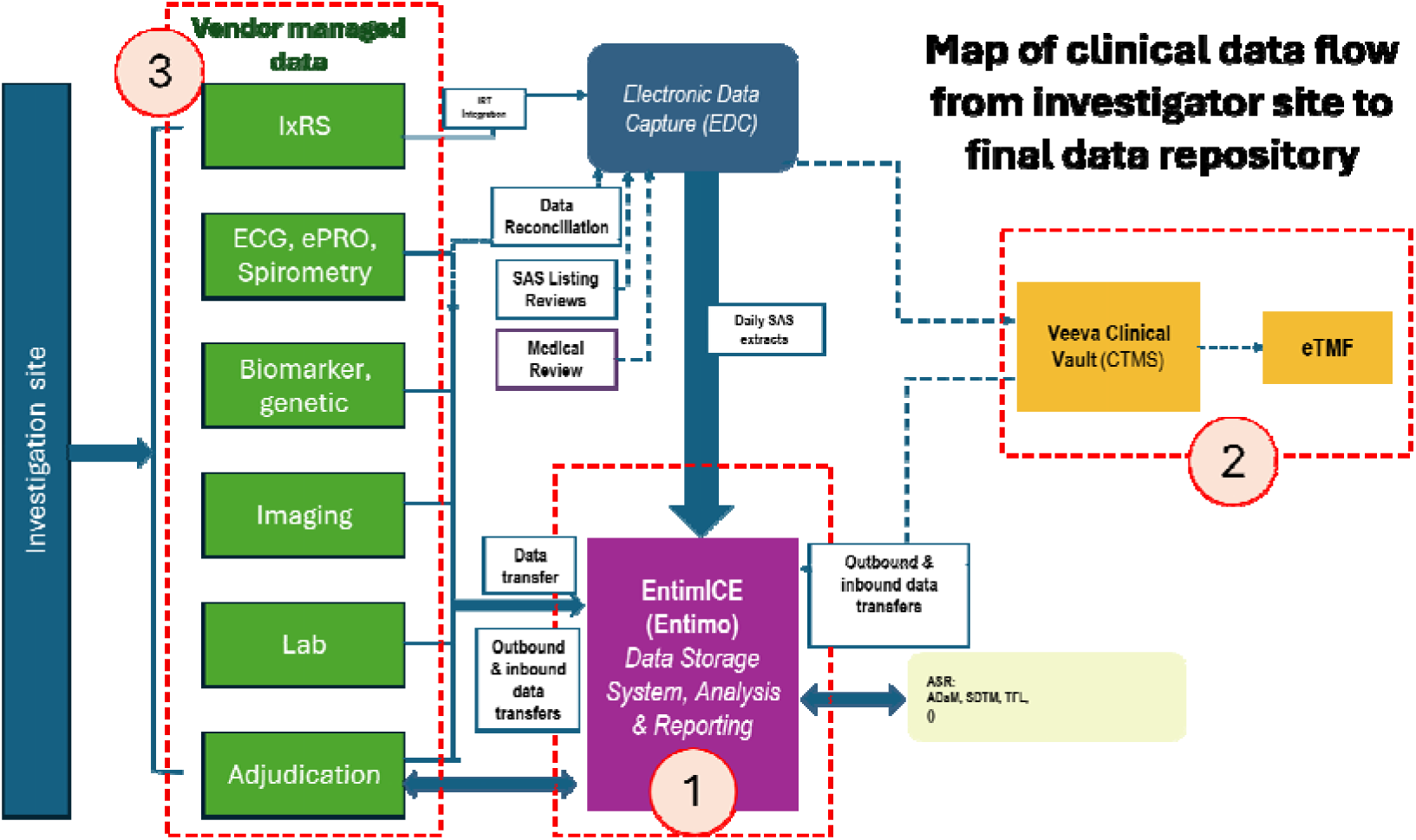
Clinical trial data flow schematic. 1. The EntimICE system (purple box) receives distilled trial-critical numeric data for analysis. 2. The final TMFs are stored in the Veeva Clinical Vault (yellow box). 3. The different data streams for a typical trial, which are generated at the investigator sites (green boxes). Different vendors are involved in collecting, storing, and processing this data.

## Methods

Based on the trial data flow described above (Figure 1), three potential emission hotspots in the trial data storage were identified. These hotspots were determined based on the volume of data flowing through them. This includes (i) data processing and storage in the EntimICE statistical analysis environment, (ii) the eTMFs storage in Veeva Clinical Vault (VCV), and (iii) CRO-managed raw and cleaned trial data.

### Study window

This study was conducted between April 2024 and September 2025. The assessment window for the clinical trials considered in the study, i.e., DAPA-HF and ADRIATIC, which were termed the ‘active period’ of the study, spanned the commencement of the trial to the completion of data analysis. International regulatory guidelines such as ICH-GCP, and AZ’s data retention policy recommend archiving clinical trial data for up to 25 years after study completion. The Discussion section addresses the potential emissions associated with this long-term data archiving.

### Emissions from statistical data analysis

There are two components to the EntimICE system: (i) computational power used for data analysis, and (ii) storage of active and recently completed trial data needed for the analyses. For the first component, the number of data servers and the average power usage per hour (Watt-hour) for each server were determined based on discussions with the EntimICE technical engineers and product owners^12^. The CO_2_ emissions per kWh of electricity used were determined using a standard geo-localised grid electricity emission factor from Electricity Maps^13^. The average unit emission for ten days between February 2025 and March 2025, during which the calculations were conducted, was used.

For the second component, the number of files, total file size in gigabytes (GB), and the estimated total size of the stored active trial files across all trials housed in the EntimICE system were calculated to assess per-GB power utilisation^14^. The power utilisation for storing 1GB of data was determined using established conversion factors, i.e. for data storage, 1 GB = 0.0046 kWh^15^.

The power utilisation for the other operations such as networking and server cooling, was not accounted for in this calculation, because the server site also hosted other AZ operation servers; hence, it was infeasible to isolate the power utilisation specifically for the EntimICE server operations. This remains a significant limitation of this study.

In addition, two representative AZ trials, DAPA-HF^16^ and ADRIATIC^17^, were chosen to determine the number of files and total file size for each trial. This was done to estimate the per-trial analytical data file size and to assess whether trial size and complexity affected it. Furthermore, the overall CO_2_ emissions from these two trials were previously reported^1^

### Emissions from TMF storage

The second emission hotspot that was considered for emission calculations was the file size and location of the electronic TMFs (eTMFs). The eTMFs for AZ’s ongoing and completed clinical trials are stored in the Veeva Clinical Vault (VCV), a product offered by Veeva. Veeva stores data across a combination of on-premises and hyperscale cloud servers. Quantifying trial-specific emissions from eTMF storage was not feasible because Veeva’s database structure does not allow emissions to be assessed at the trial level. However, we were able to determine the total size of all Veeva-held TMFs across current and archived AZ studies.

### Emissions from upstream trial data stored by third-party clinical research organisations (CROs)

Two of the largest CRO partners for AZ’s clinical development were identified and were provided with an information requisition questionnaire (Table S1). For CRO-held data, since the data is stored both on-premises and on third-party cloud servers, the specific server location was not captured. Only the volumes of data in terms of the total number of trials and the total file sizes were collected. This was used to estimate the total volume of data held by the two vendors for AZ-sponsored trials.

## Results

### Emissions from data analysis in the EntimICE system

The EntimICE® system is an integrated clinical data analysis enterprise software suite offered by the company Entimo^18^. It was the central statistical analysis environment at AZ and is widely used by other clinical development organisations. The majority of AZ’s trial-related statistical analyses are performed using the EntimICE system. It consists of a metadata-driven Statistical Computing Environment (SCE) embedded in the Clinical Data Repository and the Metadata Repository ^19^.

The servers that hold AZ’s EntimICE system are based in Sweden. The AZ EntimICE system consists of 4 SAS job servers, in addition to other DB, FAS and FTP servers. For this study, only the computational power of the four SAS servers was calculated. The other database infrastructure and servers also served other AZ divisions, and the emissions attributable to clinical trial activity were assessed as negligible.

The annual average power consumption and the maximum allocated power for each of the four servers are shown in Table S2. The stepwise calculation to arrive at the annual emissions is shown in Table 1. Based on the average and maximum power usage (A), the daily (B) and annual power consumption (D) were calculated. Using this value, kg CO_2_eq were calculated using the emission conversion rate for Sweden (20 gCO_2_eq/kWh)^20^. The conversion was cross-verified using a standard online conversion calculator^21^.

**Table 1.** Emission calculation based on average usage.

| Server Name | Average Usage (W) | Power consumption if the servers are used for 24 hours | Converting to KWh | kW consumption per year | KgCO<sub>2</sub>eq emissions per year (kg CO<sub>2</sub> eq) |
| --- | --- | --- | --- | --- | --- |
| Calculation | 'A' | 'B' = (A*24) | 'C' = B/1000 | 'D' = C*365 | 'E' = D*0.020 |
| Server 1 | 327 | 7848 | 7.84 | 2864.52 | 57.29 |
| Server 2 | 252 | 6048 | 6.04 | 2207.52 | 44.15 |
| Server 3 | 240 | 5760 | 5.76 | 2102.4 | 42.05 |
| Server 4 | 255 | 6120 | 6.12 | 2233.8 | 44.68 |
| Maximum power | 550 | 13200 | 13.2 | 4818 | 96.36 |

The emissions from clinical trial data analysis across all active trials at AZ, using the four SAS servers, range from 42.05 kg CO_2_eq to 96.36 kg CO_2_eq. The assumptions used in this calculation were that the four servers were used 24 hours a day, 365 days a year, at average and maximum allocated power utilisation. However, in general practice, the servers are unlikely to meet the maximum power allowance.

### Emissions from trial-level data storage for analyses using EntimICE

To determine the trial-level emissions associated with data analysis, the DAPA-HF and ADRIATIC studies were used as examples. The number of trial-related folders and the folder sizes are presented in Table S3 and Table S4. The conversion factor used for data storage (0.0046 kWh / GB) has been previously reported^4^.

Using this conversion factor, storing all the DAPA-HF files and ADRIATIC study files on EntimICE for a year would result in approximately 209 kg CO_2_eq and 636 kg CO_2_eq per annum, respectively (Table 2). The study data is stored in EntimICE for analysis for the duration of the study. Therefore, for ADRIATIC, which started in 2018 and is expected to be completed in 2026, the emissions from data analysis are expected to be 5088 kg CO_2_eq.

**Table 2.** Emission calculation for data storage on EntimICE servers.

| Data | Total data (GB) | Total power<br>consumption per<br>day (kWh) | Total power<br>consumption per<br>year (kW) | Total<br>emission per<br>year<br>(kg CO<sub>2</sub>eq) |
| --- | --- | --- | --- | --- |
| Calculation | 'A' | 'B' = A*24 *<br>0.0046 | 'C' = B*365 | D' = C*0.20 |
| DAPA-HF | 260.19 | 28.7249 | 10484.62 | 209.69 |
| ADRIATIC | 789.7 | 87.1828 | 31821.75 | 636.43 |
| For 100 TB of<br>data storage | 100000 | 11040 | 4029600 | 80592 |

The EntimICE system holds 100-125 TB of clinical trial data across all active and recently completed AZ clinical trials, equating to ∼80.5 to 100 tonnes of CO_2_eq emitted per year.

### Emissions from the final eTMF storage

We determined the file sizes stored in the Veeva Clinical Vault (VCV) (Table 3). As explained in the methods, we were unable to calculate direct emissions due to the database architecture and because Veeva’s data is partially stored on external hyperscale data centres. However, based on publicly available third-party-derived information regarding emissions from data storage, and given the file sizes (a few 100 GBs per trial), the emissions associated with bulk storage of these files are expected to be negligible^22^.

**Table 3.** eTMF file sizes stored in the VCV.

| Study | Size of file |
| --- | --- |
| DAPA-HF | 110.97 GB |
| ADRIATIC | 107.57 GB |
| All active studies | 9.63 TB |
| All archived studies | 1.87 TB |

### Emissions from upstream trial data stored by clinical trial vendors

The third component assessed was the volume of clinical trial data stored upstream by third-party CROs managing AZ’s clinical trials. As explained earlier, the green boxes in Figure 1 illustrate different data components held by vendors. A large portion of the trial data generated at the investigator sites is stored by the various suppliers responsible for the different protocol-defined data points. The suppliers maintain the raw files and the cleaned data and shared for data analysis.

We approached two of the largest AZ vendors (vendors A and B) by volume of business and provided them with a questionnaire (Table S1) to assess the volume of clinical trial data from AZ-sponsored studies they held in their data servers. The summary of the trial data held by the two vendors is shown in Table 4. As the data is stored in the database structure and there are multiple sub-components, such as recruitment information, lab data, payments, eConsent, etc., that are accessed individually. This architecture does not segment data by trial at the storage level, making it technically infeasible to isolate and quantify a single trial-specific data footprint.

**Table 4.** Vendor-held trial data.

| Vendor | Total archived trials | Total active trials | Total file data size |
| --- | --- | --- | --- |
| Vendor A | 850 | 136 | ~101 TB |
| Vendor B | 64 | 247 | ~700 GB |

It was hypothesised that the storage and processing of imaging data are likely to have the greatest impact on emissions due to the large file sizes of individual patient data. Therefore, for the last component of this study, we focused on the volume of imaging data specifically. Imaging vendors are involved with AZ from trial start-up through project planning and partner with the investigator sites to collect imaging data. Clinical data images are collected, verified for accuracy, prepared for analysis, and reviewed to determine whether they meet quality standards. The analysed output is shared with AZ.

As a representative trial, we chose the ADRIATIC study to determine the total size of the imaging data. We identified the image processing vendor who was involved in the ADRIATIC study (Vendor C). We calculated that from the study’s commencement in 2018 to March 2025, ∼1.1 TB of data had been stored for the ADRIATIC study (Table 5). However, the emissions associated specifically with holding this data were not quantifiable due to data architecture constraints explained above.

**Table 5.** ADRIATIC study data stored by perceptive.

| ADRIATIC study data with Vendor C |  |
| --- | --- |
| Total active sites | 172 |
| Total subjects | 729 |
| Total timepoints (till date) | 7059 |
| Size of total data stored | ~1.1 TB |

In conclusion, regardless of the complexities involved in calculating per-trial data storage emissions, the assessment indicated that the total volume of data generated and, consequently, its associated emissions are remarkably small compared to previously published trial emissions hotspots^14^.

## Discussion

In this study, we assessed the CO_2_ emissions from clinical trial data storage and analysis. Based on a mapping exercise to track the flow of data from the investigator site to the final trial eTMF repository, three hotspots were determined for assessment: (i) data analysis and storage in the EntimICE statistical analysis environment, (ii) trial eTMFs storage in VCV, and (iii) data stored by third-party CROs. Our study highlighted the complexity involved in calculating trial-specific emissions from data generation and storage due to the nature of database architecture, multiple vendors involved in storing and managing the data, and the lack of mechanisms to accurately assess emissions from data storage.

Notwithstanding the complexity of the assessment, this study indicated that emissions from clinical trial data storage and management did not materially affect the overall CO_2_ emissions from clinical trials. For example, in the previous reports of the complete DAPA-HF study, the per-randomised-patient emissions were 527 Kg of CO_2_eq^1^. Our study now estimates data-related emissions of 209.69 Kg of CO_2_eq per year for the whole trial, which equals ∼44.2 g of CO_2_eq per patient per year during the active period of the trial.

### Impact of the nature of analyses and the server’s geographical location on emissions

Emissions from data analysis in the EntimICE statistical analysis environment were around 80 tonnes CO_2_eq per year per 100 TB of data. Of the three components, only the statistical analysis environment was entirely housed on AZ’s premises, allowing us to measure emissions with greater accuracy. The emissions from statistical analysis were minimal primarily because standard SAS-based statistical analysis using numeric data is not computationally intensive, compared to complex calculations such as genome-wide association analysis and image processing^23^. In addition, Sweden’s low-carbon-intensity electricity grid (∼20 g CO2eq per kWh) contributed to the low emissions observed in the calculations. The emissions from data centres are country-specific; the associated emissions would have been proportionally higher if the data centres were powered by electricity from a carbon-intensive grid.

### Complexities involved in estimating emissions from hyperscale data servers

Hyperscale data servers used by third-party CROs posed greater complexity in assessing per-trial emissions when compared to in-house servers, because the data was hosted across a mix of self-owned and hyperscale cloud servers. This created an insurmountable challenge in delineating and assessing per-trial emissions, because (i) the flat trial data files are converted to database components when stored in the cloud servers, the trial-specific data are coalesced with other data and become highly fragmented, and cloud servers hold data in different geographies, and each service provider uses different emission offset mechanisms. These issues require coordination across R&D and the technology and infrastructure management partners to identify key areas for emission reduction.

### Data emissions assessment window for this study and emissions from long-term data archival

Regarding the duration of trial data storage, AZ’s internal policy, and the EU Clinical Trials regulations require that trial data be stored by the sponsor for 15–25 years^24^. However, once the study is completed, the data can be moved to low-powered ‘cold storage.’ Therefore, the life of clinical trial data could be divided into two phases: ‘hot’ storage during the active phase of the study, when the data is routinely accessed for routine analyses, and long-term archival in cold storage, where it is held in line with regulatory requirements. The focus of this study was the emissions from the active hot-storage phase of the clinical trial data, as the latter was considered a low-power-consuming activity and, by extension, low-emission operation. That said, with emerging artificial intelligence (AI)-enabled trials, where large language models require large training data sets, transfer to cold storage may be considerably delayed, resulting in extended time in ‘hot’ storage.

### Future perspective

While this study showed that clinical trial data storage and analysis were not energy-intensive and therefore the associated emissions were low, this scenario may change with the integration of AI in clinical operations and data processing^25^, and with greater adoption of genomic and proteomic data analysis in clinical trials. Clinical trials routinely collect biosamples to build biological databases and biobanks. This substantially increases the need for computational resources to analyse the data. The emissions associated with such genomic analyses and molecular simulations have been shown to be orders of magnitude higher than standard statistical analyses of clinical data^26^.

### Strengths and Limitations of this study

The strengths of this study include being the first we are aware of to map the journey of trial data from the investigator site to the final data repository and to identify the emissions associated with key hotspots. The results of this study indicate that trial data management is not currently a significant contributor to the overall emissions associated with clinical trials. However, this may change as clinical trials adopt increased digital data-collection methods and the volume of data collected increases. Future clinical trial carbon emission assessment frameworks should consider including this component in their analyses.

This study has notable limitations that need to be considered when interpreting the results. Firstly, energy-consuming components beyond data storage that make up a typical data server, such as backup servers, networking, and cooling systems, were not considered because isolating their functions for clinical trial data analyses was infeasible. Secondly, due to the complexity of cloud data server infrastructure, confidentiality concerns regarding server infrastructure, and gaps in publicly available data, we did not have access to numerous data points, such as server locations, server architecture, and server emissions certifications. The availability of these data points would have improved the accuracy of our calculations.

### Conclusion

Various industry and academic groups who reported clinical trial emission calculation methodologies that use a combination of curated databases and questionnaire-based approaches^5^. None have explicitly incorporated emissions from data storage and analysis, especially in centralised data servers, into their framework. This study proposes a framework for assessing emissions from clinical trial data storage and management that utilises centralised data servers. While this study was conducted at one pharmaceutical sponsor, the results are likely relevant to most sponsors. Based on our assessment, emissions from data storage and management were minimal and thus are not a key area for trialists when calculating and reducing carbon emissions from clinical trials. However, with greater adoption of decentralised trial designs, genomic analyses within trials, and the integration of AI into trial data management, execution and analyses, the emissions associated with trial data storage and analysis may become a material component of overall trial emissions. We encourage other groups to test our framework, fine-tune it for their trial-specific data storage and analysis systems, and identify hotspots that we may have missed in our study.

## Acknowledgements

We thank AstraZeneca colleagues, especially Jen Valsler and Maria Hedwall, for their guidance and strategic input on this work. We thank the CRO vendors for their support. We thank Loïc Lannelongue, Assistant Research Professor at the University of Cambridge, for his insights on sustainable computing practices.

## Author Contributions

The study was conceptualized and executed by HSP and NM. HSP was the primary author of the manuscript with guidance from NM.

## Ethics statements

Patient consent for publication: Not applicable.

## Ethics approval

This study does not involve human participants. The trials appraised for their carbon footprint in this study were approved by the ethics committee at each site. All participants provided written informed consent.

## Data availability statement

All data relevant to the study are included in the article or uploaded as online supplemental information. Data obtained from a third party may not be freely publicly available. Financial and third-party data related to some of the findings described in this manuscript may be considered commercial in confidence and not publicly available. Data underlying the findings described in this manuscript may be obtained in accordance with AstraZeneca’s data sharing policy described at https://astrazenecagrouptrials.pharmacm.com/ST/Submission/Disclosure.

## References

1. Mackillop N, Shah J, Collins M, et al. Carbon footprint of industry-sponsored late-stage clinical trials. BMJ Open 2023;13(8):e072491. doi: 10.1136/bmjopen-2023-072491

2. Griffiths J, Fox L, Williamson PR. Quantifying the carbon footprint of clinical trials: guidance development and case studies. BMJ Open 2024;14(1):e075755. doi: 10.1136/bmjopen-2023-075755

3. LaRoche JK, Lanier J, Alvarenga R, et al. Climate footprint of industry-sponsored in-human clinical trials: life cycle assessments of clinical trials spanning multiple phases and disease areas. BMJ open 2025;15(2):e085364.

4. Griffiths J, Adshead F, Salman RA-S, et al. What is the carbon footprint of academic clinical trials? A study of hotspots in 10 trials. BMJ open 2024;14(10):e088600.

5. Keegan D, Brennan L, O’Neill L, et al. Measuring the carbon footprint of clinical research activities: a scoping review of measurement tools and methods. BMJ Open 2026;16(8):e119989. doi: 10.1136/bmjopen-2026-119989

6. LaRoche JK, Alvarenga R, Collins M, et al. Climate footprint of industry-sponsored clinical research: an analysis of a phase-1 randomised clinical study and discussion of opportunities to reduce its impact. BMJ Open 2024;14(1):e077129. doi: 10.1136/bmjopen-2023-077129

7. Cranley D, Dunn S, Taylor JP, et al. Carbon footprint of a sample of clinical trials for people with neurological disorders: cross-sectional analysis. BMJ Open 2025;15(6):e090419. doi: 10.1136/bmjopen-2024-090419 [published Online First: 2025/06/18]

8. Fragão-Marques M, Ozben T. Digital transformation and sustainability in healthcare and clinical laboratories. Clin Chem Lab Med 2023;61(4):627–33. doi: 10.1515/cclm-2022-1092 [published Online First: 20221206]

9. ClinicalTrials.gov: A 25-Year Journey to a Half-Million Registered Studies. NLM’s ClinicalTrialsgov, 2025.

10. The 5Vs of Clinical Data. In: (SCDM) SoCDM, ed., 2022.

11. Deb T. Clinical Data Management System Market to Reach USD 9.8 Billion by 2034 at 11.2% CAGR 2026 [Available from: https://media.market.us/global-clinical-data-management-system-market-news/.

12. GITC-Engineers. AstraZeneca plans to invest Rs 250 crore to grow its Global Innovation and Technology Centre in India, bringing some 1,300 highly skilled roles by 2025, 2024.

13. Electricity-Maps. Live 24/7 CO emissions of electricity consumption | App | Electricity Maps 2025 [Available from: https://app.electricitymaps.com/map/72h/hourly accessed July 2025.

14. EntimICE-AZ. EntimICE system Owner, 2024.

15. LCCT-Calculator. LCCT Calculator 2024 [Available from: https://clinicaltrialcarbon.org/ accessed July 2025.

16. McMurray JJV, Solomon SD, Inzucchi SE, et al. Dapagliflozin in Patients with Heart Failure and Reduced Ejection Fraction. New England Journal of Medicine 2019;381(21):1995–2008. doi: doi:10.1056/NEJMoa1911303

17. Cheng Y, Spigel DR, Cho BC, et al. Durvalumab after Chemoradiotherapy in Limited-Stage Small-Cell Lung Cancer. New England Journal of Medicine 2024;391(14):1313–27. doi: doi:10.1056/NEJMoa2404873

18. Entimo. entimICE DARE - Statistical Computing Environment 2025 [Available from: https://www.entimo.com/entimice-dare accessed July 2025.

19. PhUSE 2016 The development of standards management using EntimICE-AZ. Pharmaceutical Users Software Exchange 2016; 2016. Lex Jansen.

20. Nowtricity [Available from: https://www.nowtricity.com/country/sweden/ accessed April 2025.

21. Cencepower [Available from: https://www.cencepower.com/calculators/kwh-to-co2-calculator accessed April 2025.

22. Climatiq. Emission Factor: AWS (us-east-1) CPU | Information and Communication | Cloud Computing - CPU | Virginia, US | Climatiq 2025 [Available from: https://www.climatiq.io/data/emission-factor/e8fc4ce0-8013-48ea-a86c-c40f73bf1da9 accessed July 2025.

23. Lannelongue L, Grealey J, Bateman A, et al. Ten simple rules to make your computing more environmentally sustainable. PLOS Computational Biology 2021;17(9):e1009324. doi: 10.1371/journal.pcbi.1009324

24. Regulation (EU) No 536/2014 of the European Parliament and of the Council: https://www.legislation.gov.uk/; 2014 [Available from: https://www.legislation.gov.uk/eur/2014/536/article/58 accessed August 2026.

25. Aczel M, Chamanara, S., Matin, M., Farsi, A., Marwala, T., Madani, K. Environmental Cost of AI’s Energy Use: Carbon, Water and Land Footprints. In: United Nations University Institute for Water EaHU-I, ed. UUNU-INWEH Report, 2026.

26. Grealey J, Lannelongue L, Saw W-Y, et al. The Carbon Footprint of Bioinformatics. Molecular Biology and Evolution 2022;39(3) doi: 10.1093/molbev/msac034

