## Supplementary document for "Mapping the flow of digital data in clinical trials and assessing its carbon footprint" (from another version of this preprint: version 1, 2026-08-03)

**Supplementary data**

Table S 1 CRO - Clinical data volume assessment - secondary information request questionnaire

| Active trials: | |
| --- | --- |
| Question | Description |
| 1. How many active AstraZeneca clinical trials are you currently involved in, across therapy areas? | Please provide the number of AstraZeneca active clinical trials that you are currently involved in. |
| Ans. | E.g., 9 |
| 2. Please provide a breakdown of the of trails by therapy area: | Please give the number of active trials in each therapy area |
| Ans.  ONC + Haem:  Respiratory & Immunology:  CVRM:  Vaccines and Immune therapies:  Rare Disease:  Other: | E.g.,  Oncology + Haematology: **7**  Respiratory & Immunology: 0  CVRM: **2**  Vaccines and Immune therapies: 0  Rare Disease: 0  Other: 0 |
| 3. What is the total size of data (in GBs) generated so far across all these trials? | Please provide the total size of all the data files held across all the trials – in GBs or TBs |
| Ans. | E.g., 254 TB (across 9 trials) |
| 4. Please provide a breakdown by therapy area and number of trials | We are looking for the total file size, across all the trial-related files and documents. |
| Ans. Therapy Area \| No. of trials \| No. of patients \| Total size (GBs); ONC. + Haem: \| \| \|; R&I: \| \| \|; CVRM: \| \| \|; V&IT: \| \| \|; Rare Disease: \| \| \|; Other: \| \| \| | E.g., Therapy Area \| No. of trials \| No. of patients (total) \| Total size (GBs); ONC+ Haem: \| 7 \| 3250 \| 200 TB; R&I: \| 0 \| \|; CVRM: \| 2 \| 8000 \| 54 TB; V&IT: \| 0 \| \|; Rare Disease: \| 0 \| \|; Other: \| 0 \| \| |
| 5. Where are the data from the active trials currently stored? | Please provide the breakdown of how and where the data are stored |
| Ans.  On-premises data servers:  Third party cloud servers:  Both: | E.g.,  All the data are stored on AWS servers |
| 6. What is the geographic location of the servers used for data storage? | We are looking for a broad understanding of where the server might be located. Country and broad regional level (only for US) is sufficient. |
| Ans.   a) On-premises server: (Please specify country) __________ b) Cloud server: ___________ | E.g.,   1. On-premises server: (Please specify country): Germany 2. Cloud server: (Location of server of the cloud service provide): US- East |
| Concluded trials | |
| 7. How many concluded trials are you currently retaining the data for? | Please provide the number of archived, non-active AstraZeneca trials, for which you hold the data |
| Ans. Trial name / identifier \| Total size (GBs) \| Year of archival | E.g., Trial name/identifier \| Total size (GBs) \| Year of archival; DAPA-HF \| 200 TB \| 2019 |
| 8. Where are the data from concluded trials archived? | Please provide the total number of archived trials, and archived file size, based on server type |
| Ans. Server type (on prem/ cloud) \| Number of trials \| Total size (GB/TB) \| Location | E.g., Server type (on prem/ cloud service provider) \| Number of trials \| Total size (GB/TB) \| Location; On – Prem \| 0 \| \|; AWS \| 9 \| 300 TB \| US-East |

Table S 2 Average and maximum energy consumption of EntimICE servers

| **Server Name** | **Average Usage(W)** | **Allocated(W)** | **Maximum Power(W)** |
| --- | --- | --- | --- |
| Server 1 | 327 | 625 | 550 |
| Server 2 | 252 | 595 | 550 |
| Server 3 | 240 | 670 | 550 |
| Server 4 | 255 | 640 | 550 |

Table S 3 Number of files and file size for DAPA-HF study

| Master Folder | Size (GB) | Number of subfolders | Number of files |
| --- | --- | --- | --- |
| AR | 218 | 2773 | 44993 |
| DM | 37 | 502 | 5876 |
| Expl_analysis | 1.6 | 34 | 234 |
| Expl_biostat | 0.29 (296M) | 91 | 310 |
| Common | 3.3 | 42 | 294 |
| Total | 260.19 GB |  |  |

Table S 4 Number of files and file size for ADRIATIC study

| Master Folder | Size (GB) | Number of subfolders | Number of files |
| --- | --- | --- | --- |
| AR | 721 GB | 8393 | 72532 |
| DM | 66 GB | 609 | 9676 |
| doc | 0.1 GB | 8 | 390 |
| Expl | 2.5 GB | 138 | 2960 |
| Common | 0 GB (4 KB only) | 1 | 0 |
| sandbox | 0.1 GB | 142 | 40 |
| Total | 789.7 |  |  |
